# Bladder Cancer in the United States: National Trends and State-Level Patterns from Global Burden of Disease (GBD) Study, 1980-2021

**DOI:** 10.64898/2026.07.11.26357810

**Authors:** Alireza Sadeghi, Fatemeh Nouri, Niloofar Dehdari Ebrahimi, Ehsan Taherifard, Mohammadreza Soltani, Stephen B. Williams, Wassim Kassouf

## Abstract

**Background:** The U.S. has the highest health care expenditure globally. Examining long-term national and state-level trends, and benchmarking them against other health systems, can provide important insights for public health planning and policy development.

**Aim:** This study aimed to characterize temporal trends in the burden of bladder cancer, including prevalence, incidence, mortality, disability-adjusted life years (DALYs), years lived with disability (YLDs), and years of life lost (YLLs), and to evaluate state-level disparities and potential long-term effects of health policies.

**Methodology:** Data were obtained from the Global Burden of Disease (GBD) 2021 study. Age- and sex-stratified trends were analyzed and visualized at both national and state levels across the study period. Burden estimates were additionally compared with those of other major health systems, including the European Union, countries with high socio-demographic index, and high-income settings.

**Results:** In 2021, the U.S. recorded age-standardized rates of 8.35 for YLDs (0.33 lower), 3.41 for mortality (0.11 higher), 59.80 for YLLs (6.75 higher), 100.63 for prevalence (6.61 lower), 14.69 for incidence (0.51 lower), and 68.15 for DALYs (6.42 higher) compared to 1990 records. Lowest gender discrepancies across all measures were in 2021 were observed in District of Columbia with male:female ratio of DALYs: 2.4 [1.7, 3.2], mortality: 2.4 [1.8, 3.2], incidence: 2.5 [1.8, 3.4], prevalence: 2.4 [1.8, 3.2], YLDs: 2.4 [1.4, 4.1], and YLLs: 2.4 [1.7, 3.2]. In contrast, North and South Dakota had the highest gender discrepancies: DALYs: 3.9 [3.0, 5.2], incidence: 4.1 [3.1, 5.4], prevalence: 3.9 [3.0, 5.0], and YLLs: 4.0 [3.0, 5.2] and mortality: 4.2 [3.2, 5.6] and YLDs: 3.9 [2.3, 6.3].

**Conclusion:** Bladder cancer continues to impose a substantial and uneven burden across the U.S. State-level variations, driven by environmental factors, aging populations, and healthcare access gaps, require targeted prevention and improved early detection. Future research should assess the cost-effectiveness of prioritizing prevention over late-stage treatment to optimize healthcare spending.

## 1. Introduction

Bladder cancer (BC) is the sixth most common malignancy in the U.S. (1). Among American men, it ranks as the fourth most frequently diagnosed cancer and the seventh leading cause of cancer-related mortality (2). According to recent estimates from the Surveillance, Epidemiology, and End Results (SEER) Program, approximately 84,530 new cases and 17,870 deaths are projected for 2026, representing 4.0% of all new cancer diagnoses and 2.9% of cancer-related deaths nationwide (1). Similar patterns are observed globally, with an increasing absolute burden despite declining age-standardized rates, underscoring BC as a persistent and growing public health challenge (3).

Beyond its epidemiological impact, BC imposes a substantial economic burden. Its management commonly requires lifelong surveillance and repeated therapeutic interventions, rendering it the most costly cancer to treat on a per-patient lifetime basis (4–6). In addition to direct healthcare expenditures, BC exerts profound effects on patients’ quality of life and significantly impacts the emotional, social, and practical well-being of caregivers and families (7).

Substantial geographic heterogeneity in BC incidence and outcomes has been documented across the U.S. (8–11). For instance, Maine reported an age-standardized incidence rate of 26.7 per 100,000 during 2017-2021, nearly double that observed in Hawaii (13.4) (12). Such variation likely reflects differences in environmental exposures, socioeconomic conditions, access to care, and healthcare infrastructure. Incidence rates tend to be highest in northeastern states (10), whereas mortality is disproportionately greater in Southern states (13), where higher proportions of older adults, African Americans, and rural residents may experience delayed diagnosis and reduced access to high-quality treatment (14).

Despite these observations, most prior BC research has focused on national trends or single-institution cohorts, and contemporary age-standardized, state-level comparisons remain limited. Furthermore, although the Global Burden of Disease (GBD) project produces comprehensive estimates of disability-adjusted life years (DALYs), years lived with disability (YLDs), and years of life lost (YLLs) for BC, these measures have been infrequently incorporated into disease-specific epidemiological analyses. As a result, non-fatal burden and broader health-system impacts remain underexplored. National studies indicate that men experience approximately three to four times higher BC incidence than women (15), whereas women often present with more advanced disease and have poorer survival (16); however, the geographic distribution of these sex-specific disparities has not been systematically evaluated at the state level.

To address these gaps, we use data from the GBD 2021 study, which provides internally consistent estimates of key BC burden metrics across all U.S. states. This predominantly descriptive analysis offers a comprehensive assessment of BC burden across geographic regions, time, and age groups, with the aim of informing state-level public health planning and prioritization.

## 2. Methodology

### 2.1 Study Design and Data Sources

This population-based study used data from the GBD 2021 database. The GBD provides comprehensive, internally consistent, and modeled estimates for diseases and injuries across all U.S. states, enabling standardized comparisons over time, geography, age, sex, and socioeconomic strata. These features make the GBD particularly well suited for examining disparities in BC burden at national and subnational levels (17,18). All data are publicly available through the Global Health Data Exchange (GHDx).

We analyzed BC burden across all U.S. states from 1980 to 2021. Estimates for incidence, prevalence, mortality, YLLs, YLDs, and DALYs were extracted for males, females, both sexes combined, and all age groups. For international benchmarking, corresponding data were also retrieved for the European Union, high-income countries, and high Socio-Demographic Index (SDI) regions. Both age-specific and age-standardized estimates were obtained for each metric. Results are reported as rates per 100,000 population with accompanying 95% uncertainty intervals, as generated by the GBD framework. This study adheres to the Guidelines for Accurate and Transparent Health Estimates Reporting statement.

### 2.2 Definitions

#### Bladder cancer

Defined in GBD 2021 using the International Classification of Diseases (ICD) coding system: ICD-10 codes C67-C67.9 and ICD-9 codes 188-188.9, V10.51, V16.52, and V76.3 (17,19).

#### Disability-adjusted life years (DALYs)

A summary measure of population health that quantifies the gap between current health status and an ideal standard in which the entire population lives to advanced age in full health. This metric combines YLDs and YLLs due to premature mortality.

#### Years of life lost (YLLs)

The fatal component of DALYs, reflecting the number of years lost due to death occurring before the reference life expectancy.

#### Years lived with disability (YLDs)

The non-fatal component of DALYs, representing time lived in health states less than full health, weighted by severity.

#### Socio-Demographic Index (SDI)

A composite measure developed within the GBD framework that reflects levels of development based on income per capita, average educational attainment among individuals aged 15 years or older, and fertility rates among those younger than 25 years (20). The index ranges from 0 to 1 and is used to classify countries or regions into five categories: low, low-middle, middle, high-middle, and high. High-SDI countries were included as one of the comparator groups in this study.

#### Competing countries

Country groupings considered broadly comparable to the U.S. in terms of economic development and health-system context. These included the European Union, high-income countries, and high-SDI regions.

#### Age-standardized rates

Rates standardized to the GBD global age structure using the direct method, calculated as:

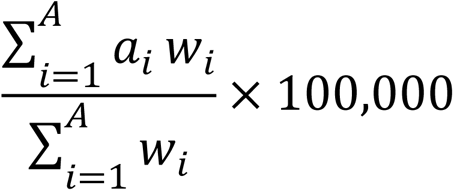

where *a_i_* denotes the age-specific rate for age group *i*, *w_i_* represents the corresponding standard population weight, and *A* is the total number of age groups.

### 2.3 Statistical Analysis

Age-standardized rates were calculated for each burden metric using the GBD 2021 world standard population. Temporal trends were evaluated across the full study period from 1980 to 2021 at both national and state levels. All rates are presented per 100,000 population with corresponding 95% uncertainty intervals.

Uncertainty Intervals (UIs) reflect the GBD Bayesian modeling framework and are defined by the 2.5th and 97.5th percentiles of 1,000 posterior draws. These intervals account for multiple sources of uncertainty, including data sparsity, measurement error, and model specification (18).

We used R v. 4.6.1 (21) and the following R packages: beepr v. 2.0 (22), flextable v. 0.9.12 (23), furrr v. 0.4.0 (24), future v. 1.70.0 (25), geofacet v. 0.2.4 (26), gganimate v. 1.0.11 (27), ggbump v. 0.1.99999 (28), ggnewscale v. 0.5.2 (29), ggtext v. 0.1.2 (30), glue v. 1.8.1 (31), knitr v. 1.51 (32–34), officer v. 0.7.5 (35), paletteer v. 1.7.0 (36), patchwork v. 1.3.2 (37), renv v. 1.2.3 (38), rmarkdown v. 2.31 (39–41), scales v. 1.4.0 (42), svglite v. 2.2.2 (43), tidyverse v. 2.0.0 (44). To enable full reproducibility, complete analytic code, data-processing workflows, and supporting files are available in the study’s GitHub repository.

#### 2.3.1 Comparison of Rates Using UIs

We calculated both absolute differences (Supporting Files) and rate ratios (Figure 5) between male and female BC burden measures. Since draw-level estimates were not available from the GBD study, uncertainty for derived quantities was approximated analytically using standard methods for independent estimates (45,46). All calculations assumed approximate normality of the log-transformed rates, which is reasonable for age-standardized rates based on large population data (47).

Let *a*_1_ and *a*_2_ represent two age-standardized rates with reported 95% UIs [*l*_1_,*u*_1_] and [*l*_2_,*u*_2_]. Standard errors were derived from the reported bounds:

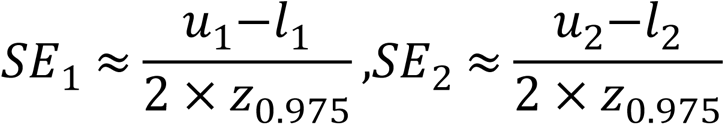

where *z*_0.975_ ≈ 1.96 is the 97.5th percentile of the standard normal distribution. The absolute difference was calculated as:

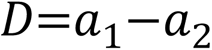

Under the assumption of independence between rates, the standard error of the difference was:

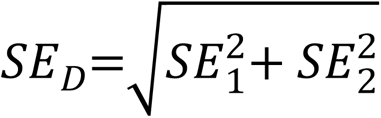

The corresponding 95% UI was then obtained as:

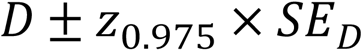

For relative comparisons, rate ratios were estimated on the logarithmic scale to account for the inherently multiplicative nature and right-skewed distribution of ratios (46). The rate ratio was defined as:

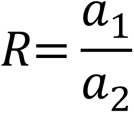

Standard errors on the log scale were approximated using the delta method:

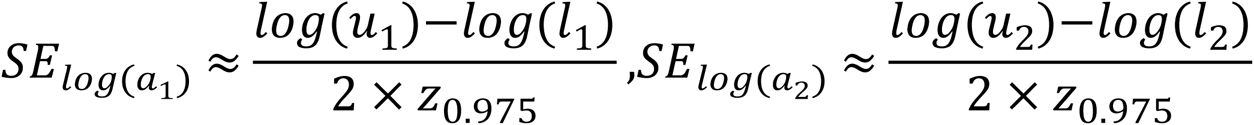

The log-transformed ratio and its standard error were calculated as:

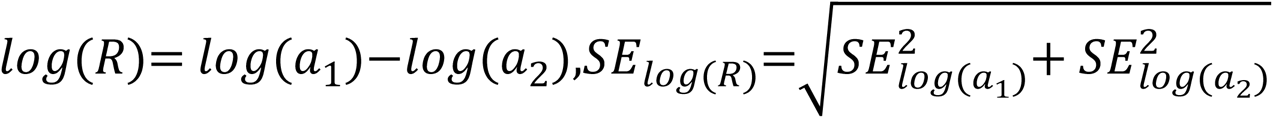

The 95% UIs for the ratio was obtained by back-transformation:

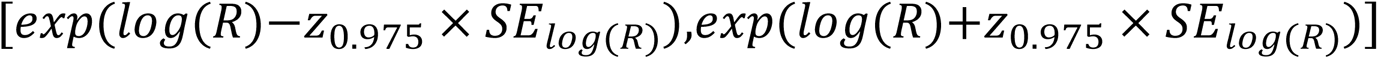

While draw-level propagation remains the preferred approach within the GBD analytical framework, the present method provides a transparent and reproducible alternative when only summary estimates are available (48). The code behind the calculations are available from the study’s Github Repository.

## 3. Results

### 3.1 U.S. Compared to Competing Countries

In 2021, the age-standardized incidence rates of BC in the U.S. were 24.0 [22.4, 25.2] and 7.1 [6.3, 7.4] for men and women, respectively, corresponding to mortality rates of 5.8 [5.2, 6.1] and 1.7 [1.4, 1.8]. In the same year, European Union, high-income, and high-SDI countries reported mortality rates of 4.6 [4.1, 4.893], 3.5 [3.1, 3.723], and 3.4 [3.0, 3.611], respectively, for both sexes combined.

With peak age-standardized incidence and prevalence rates of 16.5 [15.4, 17] and 113.9 [108.3, 117] observed in 2011, the U.S. has exhibited persistently higher incidence and prevalence of BC compared with competing countries throughout most of the 1980-2021 period. In contrast, age-standardized rates of DALYs, YLLs, and mortality in the U.S. have remained lower than those of comparator regions since the early 2000s. Notably, the pronounced and consistent declining trends observed in competing countries are largely absent in the U.S. Although these burden metrics have historically been lower in the U.S., competing countries have progressively converged toward U.S. levels over time (Figure 1).

**Figure 1.**
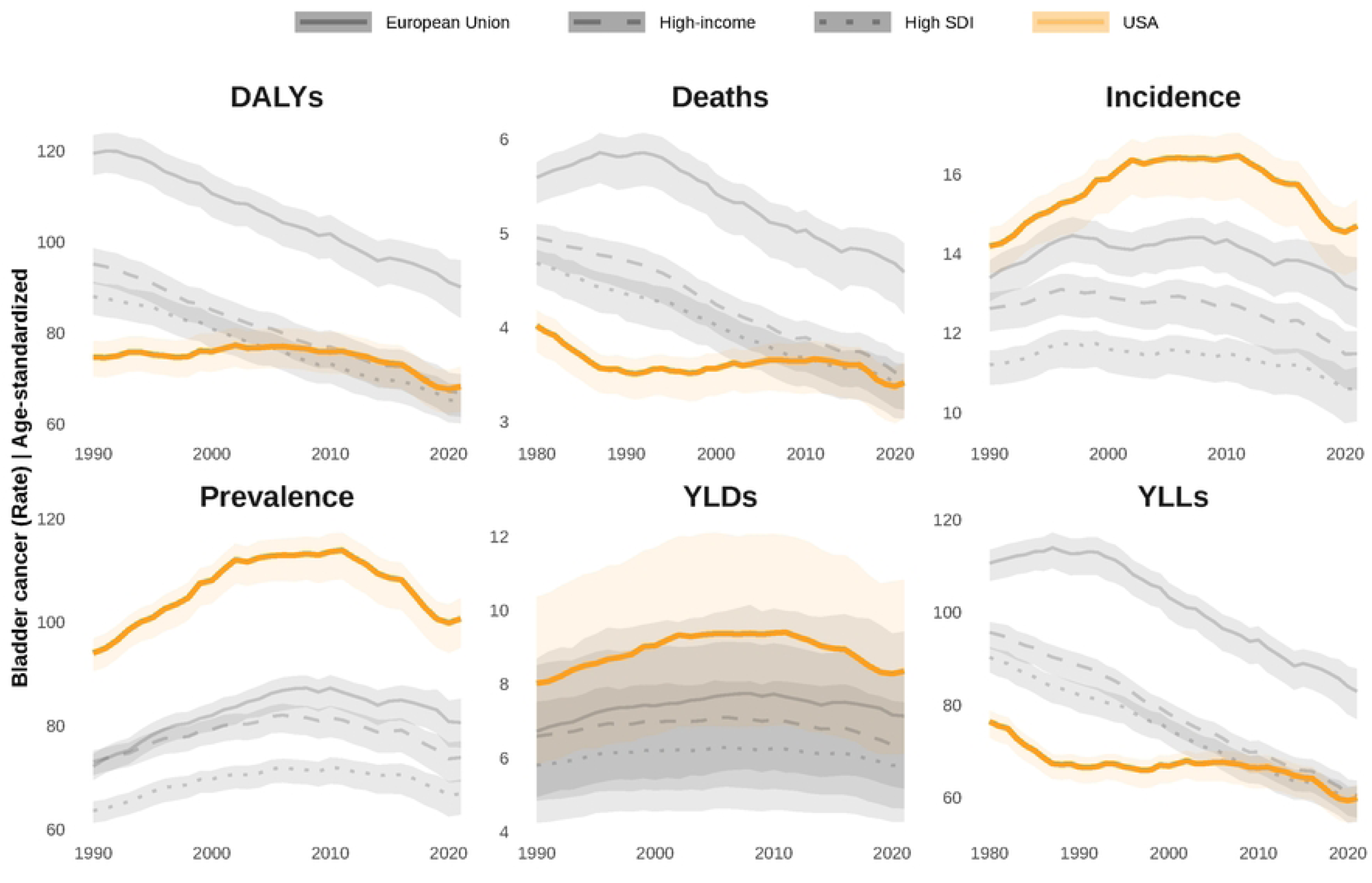
: **Age-standardized bladder cancer burden rates between 1980 and 2021 in the U.S. compared with high-income and high Socio-Demographic Index countries and the European Union.** Abbreviations: DALYs: Disability-Adjusted Life Year; YLDs: Years Lived with Disability; YLLs: Years of Life Lost

A complementary quantitative summary of the trends shown in Figure 1 is provided in Supporting Files, which presents the percentage change in age-standardized rates between 1980 and 2021 for all burden measures. Consistent with the visual trends, the table demonstrates significant reductions in DALYs (men: 34.6% [35.0%, 34.1%]; women: 26.6% [28.6%, 24.5%], men: 30.9% [31.4%, 30.5%]; women: 24.5% [26.6%, 22.3%], men: 30.2% [30.5%, 29.8%]; women: 19.4% [21.3%, 17.4%], and men: 11.8% [12.5%, 11.1%]; women: 12.4% [14.4%, 10.5%]) and YLLs (men: 32.6% [33.4%, 31.9%]; women: 26.2% [28.6%, 23.7%], men: 31.9% [32.5%, 31.2%]; women: 21.3% [23.5%, 19.2%], men: 6.7% [8.8%, 4.6%]; women: 10.5% [14.6%, 6.4%], and men: 36.4% [37.3%, 35.5%]; women: 28.4% [30.6%, 26.2%]) across high-income and high-SDI countries, the European Union, and the U.S., respectively. By contrast, while high-income and high-SDI countries and the European Union achieved significant declines in mortality (men: 30.7% [32.2%, 29.2%]; women: 26.2% [29.6%, 22.8%], men: 27.3% [28.8%, 25.8%]; women: 23.3% [27.0%, 19.7%], and men: 26.6% [27.9%, 25.3%]; women: 21.3% [24.2%, 18.4%], respectively) and male incidence (15.3% [16.4%, 14.2%], 11.8% [12.8%, 10.7%], 10.0% [10.7%, 9.4%]), changes in the U.S. were comparatively modest and statistically non-significant (0.4% [1.5%, 0.7%]).

As illustrated in Figure 2, all burden measures in 2021 were consistently higher in men across all age groups. The highest age-specific rates for each burden metric were observed in 80+ and 75-79 age strata: YLDs: 143.8 [108.0, 184.6] in men and 40.0 [28.7, 52.0] in women in 80+ years age group; Prevalence: 1,635.3 [1,526.9, 1,709.5] in men and 469.3 [415.0, 502.5] in women in 75-79 years age group; Incidence: 325.0 [274.1, 349.7] in men and 87.8 [67.6, 98.3] in women in 80+ years age group; DALYs: 1,694.0 [1,443.1, 1,829.1] in men and 480.8 [372.3, 541.0] in women in 80+ years age group; Deaths: 149.2 [125.3, 160.9] in men and 43.8 [33.8, 49.2] in women in 80+ years age group; and YLLs: 1,551.6 [1,310.0, 1,669.7] in men and 441.6 [361.3, 483.3] in women in 80+ years age group

**Figure 2.**
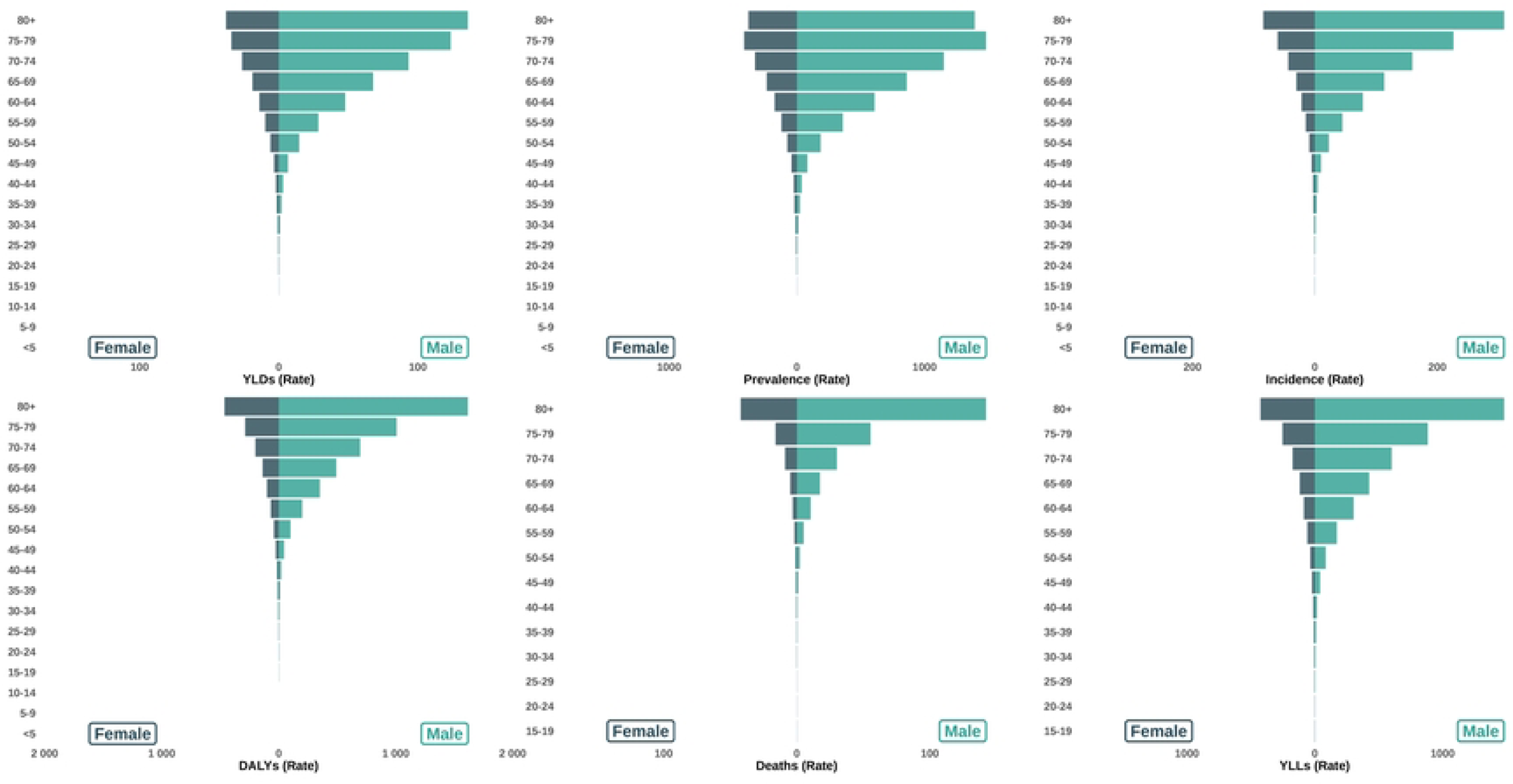
Population pyramids visualizing age-gender distribution of bladder cancer burden rates in the U.S. Abbreviations: DALYs: Disability-Adjusted Life Year; YLDs: Years Lived with Disability; YLLs: Years of Life Lost

State-level population pyramids depicting age-specific burden rates in 2021 are provided in Supporting Files and in the study’s GitHub repository. Animated population pyramids illustrating temporal changes by year are also available for each state and for the U.S. overall in Supporting Files and the GitHub repository.

In addition to age-specific snapshots, Figure 3 incorporates the temporal dimension by presenting burden rates across age groups over time. Comparable age-period vualizations for each state are available in Supporting Files and on the study’s GitHub page.

**Figure 3:**
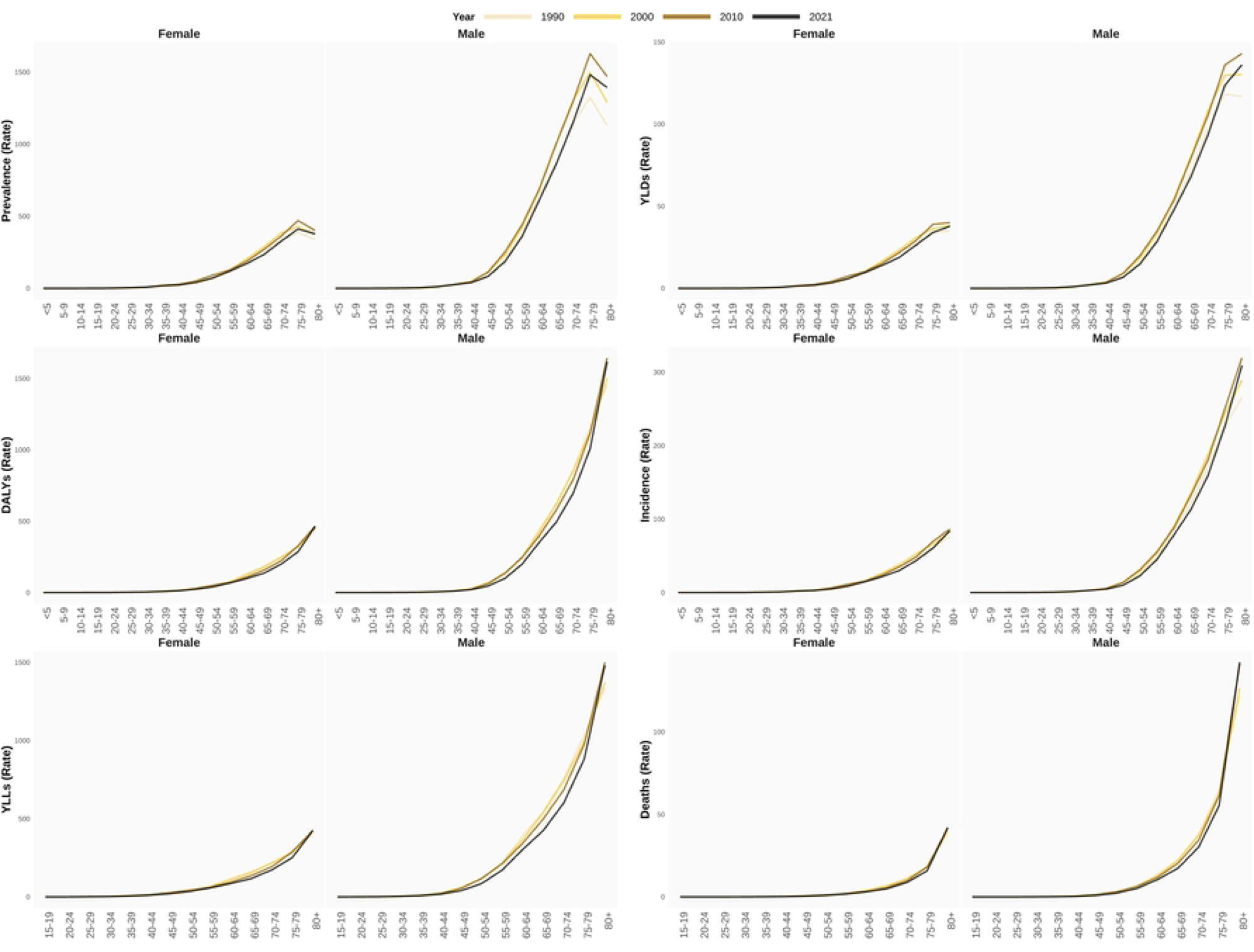
**Age-gender distribution of bladder cancer burden rates across decades in U.S.** Abbreviations: DALYs: Disability-Adjusted Life Year; YLDs: Years Lived with Disability; YLLs: Years of Life Lost

### 3.2 Bladder Cancer Burden Across the U.S

State-specific, sex-stratified temporal trends in age-standardized incidence rates are displayed in Figure 4. Analogous geofaceted plots for age-standardized prevalence, mortality, DALYs, YLLs, and YLDs are provided in Supporting Files and in the study’s GitHub repository.

**Figure 4:**
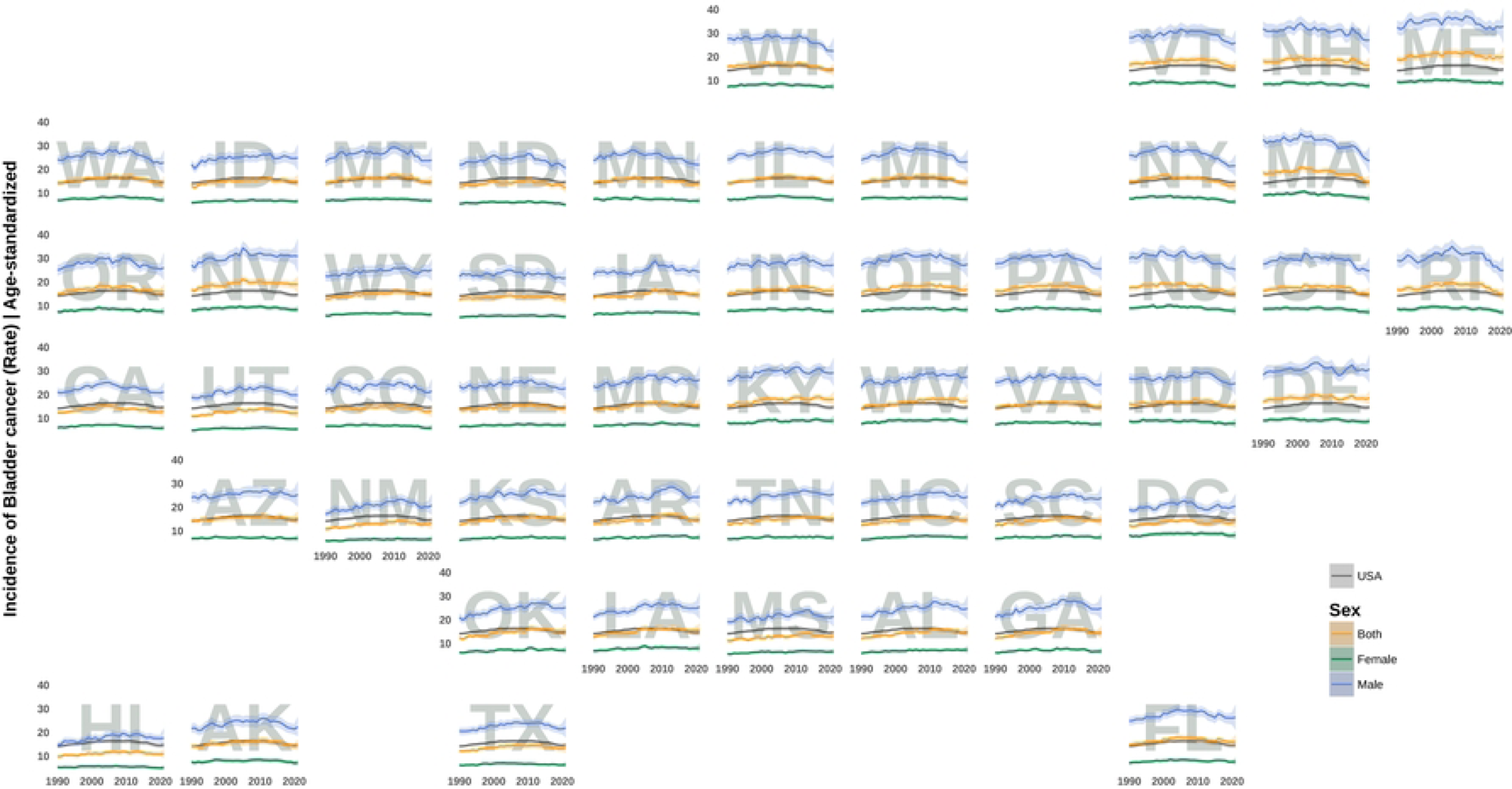
**State-level historical trends in age-standardized prevalence rates across the U.S., stratified by sex. States are arranged to approximate their geographic location.** Abbreviations: Alabama: AL; Alaska: AK; Arizona: AZ; Arkansas: AR; California: CA; Colorado: CO; Connecticut: CT; Delaware: DE; District of Columbia: DC; Florida: FL; Georgia: GA; Hawaii: HI; Idaho: ID; Illinois: IL; Indiana: IN; Iowa: IA; Kansas: KS; Kentucky: KY; Louisiana: LA; Maine: ME; Maryland: MD; Massachusetts: MA; Michigan: MI; Minnesota: MN; Mississippi: MS; Missouri: MO; Montana: MT; Nebraska: NE; Nevada: NV; New Hampshire: NH; New Jersey: NJ; New Mexico: NM; New York: NY; North Carolina: NC; North Dakota: ND; Ohio: OH; Oklahoma: OK; Oregon: OR; Pennsylvania: PA; Rhode Island: RI; South Carolina: SC; South Dakota: SD; Tennessee: TN; Texas: TX; Utah: UT; Vermont: VT; Virginia: VA; Washington: WA; West Virginia: WV; Wisconsin: WI; Wyoming: WY;

**Figure 5:**
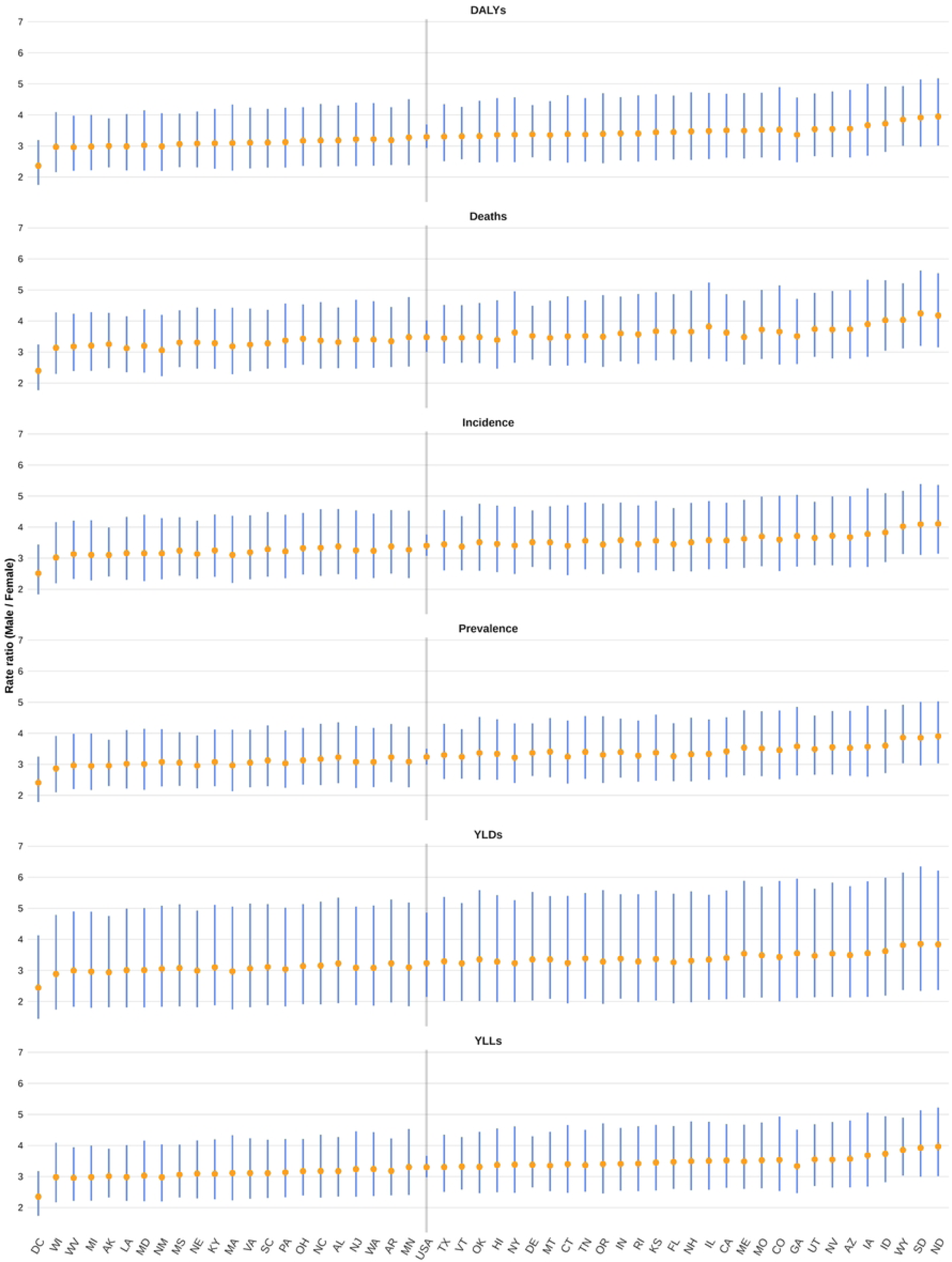
**State-level male:female ratio across different measures in 2021.** Abbreviations: Alabama: AL; Alaska: AK; Arizona: AZ; Arkansas: AR; California: CA; Colorado: CO; Connecticut: CT; Delaware: DE; District of Columbia: DC; Florida: FL; Georgia: GA; Hawaii: HI; Idaho: ID; Illinois: IL; Indiana: IN; Iowa: IA; Kansas: KS; Kentucky: KY; Louisiana: LA; Maine: ME; Maryland: MD; Massachusetts: MA; Michigan: MI; Minnesota: MN; Mississippi: MS; Missouri: MO; Montana: MT; Nebraska: NE; Nevada: NV; New Hampshire: NH; New Jersey: NJ; New Mexico: NM; New York: NY; North Carolina: NC; North Dakota: ND; Ohio: OH; Oklahoma: OK; Oregon: OR; Pennsylvania: PA; Rhode Island: RI; South Carolina: SC; South Dakota: SD; Tennessee: TN; Texas: TX; Utah: UT; Vermont: VT; Virginia: VA; Washington: WA; West Virginia: WV; Wisconsin: WI; Wyoming: WY; ; DALYs: Disability-Adjusted Life Year; YLDs: Years Lived with Disability; YLLs: Years of Life Lost

Figure 5 displays the male-to-female ratios across different states and measures in 2021 depicting a consistent gender discrepancy in the burden of BC. District of Columbia had the lowest gender discrepancies across all measures with male:female ratios of DALYs: 2.4 [1.7, 3.2], mortality: 2.4 [1.8, 3.2], incidence: 2.5 [1.8, 3.4], prevalence: 2.4 [1.8, 3.2], YLDs: 2.4 [1.4, 4.1], and YLLs: 2.4 [1.7, 3.2]. On the other hand, two states had the highest gender ratios in 2021; North Dakota with DALYs: 3.9 [3.0, 5.2], incidence: 4.1 [3.1, 5.4], prevalence: 3.9 [3.0, 5.0], and YLLs: 4.0 [3.0, 5.2] and South Dakota with mortality: 4.2 [3.2, 5.6] and YLDs: 3.9 [2.3, 6.3].

## 4. Discussion

This study used GBD 2021 data to characterize long-term trends in BC incidence, mortality, YLLs, YLDs, and DALYs in the U.S. from 1980 to 2021. At the national level, age-standardized incidence rates gradually declined beginning in the late 2010s, accompanied by a more modest decrease in age-standardized mortality rates. These improvements likely reflect advances in preventive strategies, evolving treatment strategies, and broader access to specialized care. However, national averages obscure substantial heterogeneity at the state level. While several Northeastern states experienced pronounced declines in both incidence and mortality-particularly among men-other states, including West Virginia, Kentucky, Delaware, and Nevada, showed persistently elevated or plateauing trends. Among states with consistently extreme values, Maine exhibited the highest age-standardized incidence rates throughout most of the study period, albeit with recent declines, whereas Hawaii consistently reported the lowest incidence and mortality rates Supporting Files. These contrasts likely reflect regional differences in environmental exposures, healthcare access, and the effectiveness of public health interventions.

Despite having one of the highest per-capita healthcare expenditures globally (49), the U.S. continues to report comparatively high age-standardized incidence rates of BC relative to many high-income and high-SDI regions, consistent with prior reports (3). This pattern likely reflects the country’s advanced diagnostic capacity and widespread use of cystoscopy and imaging, which facilitate more complete case ascertainment rather than a true excess in underlying disease risk (50–52). At the same time, this discrepancy highlights a potential imbalance in healthcare priorities, with disproportionate emphasis on diagnosis and treatment relative to prevention and risk reduction (53). The persistently high incidence is particularly concerning given evidence that up to 80% of BC cases diagnosed over the past two decades may be attributable to preventable exposures (54), whereas inherited genetic factors account for only a small proportion of cases in Western populations (55). Together, these findings underscore the importance of strengthening prevention-oriented public health strategies, including smoking cessation, occupational safety, and mitigation of environmental carcinogen exposure (56).

Age-specific analyses revealed a consistent increase in BC incidence, mortality, and DALYs with advancing age across all states, in line with prior literature (3). Although age-specific patterns remained largely stable over time, data from 2021 demonstrated particularly high incidence and mortality rates in the 75-80+ age groups in several populous states, including California, Texas, and Connecticut Supporting Files. This trend likely reflects demographic aging, as the U.S. population aged 65 years and older increased by 38.6% between 2010 and 2020 (56). In addition, recent evidence suggests that the oldest adults-especially men aged 85 years and older-have not experienced the same improvements in cancer mortality as younger seniors, with the largest relative increases observed in individuals aged 75-79 years and 80 years or older (57). These findings indicate that, despite overall progress, the burden of BC among older adults remains a significant public health challenge.

Consistent with prior studies, our analysis confirms that men experience BC burden rates approximately four times higher than women (15). Age-period analyses across states revealed broadly similar age-related trajectories in both sexes, characterized by a sharp rise after age 60; however, women consistently exhibited lower absolute rates and a slower decline in mortality over time. Previous studies have shown that women are more likely to present with advanced disease and have poorer survival outcomes (16,58,59). One contributing factor may be delayed evaluation of hematuria in women, as symptoms are more frequently attributed to urinary tract infections or gynecologic conditions (58,60). Addressing these disparities will require improved adherence to updated clinical guidelines, including the 2025 American Urological Association recommendations (61), alongside the incorporation of validated biomarkers for risk stratification and the adoption of sex-aware diagnostic pathways, which account for different presentations and burden rates in both genders in primary care settings.

Beyond healthcare delivery factors, emerging evidence points to biological mechanisms underlying sex-based disparities in BC. Differences in immune surveillance, hormonal milieu, hormone receptor signaling, and genetic or epigenetic alterations have been implicated (62). For example, androgen signaling may promote oncogenic pathways in men, whereas sex-specific immune responses may influence tumor behavior and progression in women (63,64). Importantly, these differences persist even after adjustment for known exposures such as smoking and occupational hazards (65). Ongoing research into sex hormones, immune checkpoints, and tumor genomics may help clarify why men are disproportionately affected by BC and why women experience worse outcomes once diagnosed.

Geographic heterogeneity observed in this study align with previous reports demonstrating the highest BC incidence among men and in Northeastern states, including Maine, New Hampshire, and Vermont (10). Current evidence has been unable to explain these patterns by factoring for smoking alone (66), which indicates that other public health factors are involved in these disparities. Environmental carcinogens, particularly arsenic contamination of drinking water from private domestic wells, have been implicated in elevated incidence in New England (66,67). Recent declines in incidence in some Northeastern states may reflect improvements in water treatment and broader public health interventions (68). For example, professionally installed and maintained household arsenic treatment systems in Maine and New Jersey have achieved substantial reductions in exposure and associated cancer risk (69), although incomplete effectiveness in a subset of systems highlights the need for continued monitoring and maintenance. Regulatory actions, such as New Hampshire’s adoption of a stricter arsenic standard for public drinking water in 2021 (70), represent important steps toward long-term risk reduction, although the long latency of BC means that benefits may take decades to fully emerge. However, interstate comparisons within the Northeast should be interpreted cautiously. States such as New York differ substantially from more rural neighboring states in terms of population density, reliance on private domestic wells, and patterns of environmental exposure, which may partially explain observed differences in temporal trends independent of policy effectiveness.

In contrast, several states in the South and West exhibit disproportionately high BC mortality despite more moderate incidence. Nevada and Southern states such as West Virginia, Kentucky, and Alabama demonstrated some of the highest age-standardized mortality rates, consistent with prior studies (13,71). Elevated mortality in these regions may reflect higher smoking-attributable cancer burdens (72), greater rurality (73,74), and reduced access to timely diagnostic and treatment services (74,75). Lower insurance coverage and longer travel distances to specialized care centers may further contribute to delayed diagnosis and worse outcomes (76). By contrast, the Northeast’s relatively low uninsured rates (77) may partially explain why states in this region experience high incidence but comparatively lower mortality, resulting in a clear regional mismatch between disease occurrence and survival.

Hawaii represents a notable outlier, with consistently low BC incidence and mortality. Several factors likely contribute to this favorable profile, including a predominantly Asian and Pacific Islander population, which has historically lower BC risk compared with non-Hispanic White populations (78), low adult smoking prevalence (79), and strong tobacco control policies (80). In addition, industrial and environmental carcinogen exposures are generally less prevalent, and access to healthcare and insurance coverage is relatively high (81). Together, these demographic, behavioral, and structural factors likely explain Hawaii’s consistently low BC burden, underscoring the potential impact of comprehensive prevention and access-to-care strategies.

The strengths of this study include the use of robust, population-based estimates to examine long-term temporal trends and state-level disparities in BC burden across the U.S. Nevertheless, our findings must be interpreted within the context of the study design. GBD estimates rely on modeled data due to incomplete or heterogeneous primary data sources, which may introduce uncertainty, particularly in states with sparse reporting. Although advanced statistical methods are used to mitigate these limitations, some degree of imprecision is unavoidable. Additionally, the inherent lag in GBD data availability means that trends beyond 2021 are not captured. Finally, this analysis is primarily descriptive and does not directly evaluate individual-level risk factors or clinical characteristics, limiting causal inference regarding the drivers of observed patterns. Most of the trends reported in this study are age-standardized; therefore, discrepancies with surveillance systems that report crude or partially adjusted rates, such as some Centers for Disease Control and Prevention (CDC) summaries, likely reflect methodological differences rather than true epidemiological disagreement. Notably, Nevada remained among the states with persistently elevated mortality trends even after age standardization.

## 5. Conclusions

BC burden in the U.S. remains unevenly distributed, with persistent state-level disparities driven by environmental, demographic, and healthcare access factors. While national trends show modest improvement, targeted interventions focusing on prevention and early detection are needed. Future economic analyses should evaluate resource reallocation from late-stage treatment to prevention to optimize healthcare spending and reduce long-term disease burden.

## Data Availability

All data are publicly available through the Global Health Data Exchange (GHDx). The complete analytic workflow, including cleaned datasets and all code required to reproduce the analyses, figures, and tables presented in this manuscript, is available in the study's GitHub repository. Data are provided in R's native binary format (RDS). Full reproducibility can be achieved by cloning the repository and rendering the project using the supplied scripts and documentation.

https://ghdx.healthdata.org/

https://github.com/alireza5969/gbd-bladder-cancer

## Supporting Files

Supporting Files are provided as appendices to the manuscript and are also publicly accessible at the links below:

1. **Percentage change between 1980 and 2021**

Percentage change in age-standardized rates between 1980 and 2021 in the U.S. compared with competing countries. Bars and error lines indicate mean percentage change and 95% confidence intervals. Green, yellow, and red denote significant decrease, non-significant change, and significant increase, respectively. This table is available from the project’s GitHub repository

2. **Geographically faceted state-level trend plots**

Geofacet plots illustrating historical age-standardized trends in bladder cancer burden measures across U.S. states are available from the project’s GitHub repository.

States are abbreviated as DALYs: Disability-Adjusted Life Year; YLDs: Years Lived with Disability; YLLs: Years of Life Lost; European Union: EU; SDI: Socio-Demographic Index; Alabama: AL; Alaska: AK; Arizona: AZ; Arkansas: AR; California: CA; Colorado: CO; Connecticut: CT; Delaware: DE; District of Columbia: DC; Florida: FL; Georgia: GA; Hawaii: HI; Idaho: ID; Illinois: IL; Indiana: IN; Iowa: IA; Kansas: KS; Kentucky: KY; Louisiana: LA; Maine: ME; Maryland: MD; Massachusetts: MA; Michigan: MI; Minnesota: MN; Mississippi: MS; Missouri: MO; Montana: MT; Nebraska: NE; Nevada: NV; New Hampshire: NH; New Jersey: NJ; New Mexico: NM; New York: NY; North Carolina: NC; North Dakota: ND; Ohio: OH; Oklahoma: OK; Oregon: OR; Pennsylvania: PA; Rhode Island: RI; South Carolina: SC; South Dakota: SD; Tennessee: TN; Texas: TX; Utah: UT; Vermont: VT; Virginia: VA; Washington: WA; West Virginia: WV; Wisconsin: WI; Wyoming: WY;

3. **State-level population pyramids** (**2021**)

Population pyramid plots depicting age- and sex-stratified bladder cancer burden for each U.S. state in 2021 are available from the project’s GitHub repository.

4. **Animated population pyramids**

Animated population pyramid plots visualizing temporal changes in age- and sex-stratified bladder cancer burden for each U.S. state are available from the project’s GitHub repository.

5. **State ranking bump plots**

Bump plots ranking U.S. states according to bladder cancer burden metrics in 1990, 2000, 2010, and 2021 are available from the project’s GitHub repository.

6. **Age-period trend plots**

Line plots illustrating age- and sex-stratified bladder cancer burden across calendar years for each U.S. state are available from the project’s GitHub repository.

## List of abbreviations

BC: bladder cancer
SEER: Surveillance, Epidemiology, and End Results
GBD: Global Burden of Disease
DALYs: disability-adjusted life years
YLDs: years lived with disability
YLLs: years of life lost
GHDx: Global Health Data Exchange
SDI: Socio-Demographic Index
UIs: uncertainty intervals
SE: standard error
AL: Alabama
AK: Alaska
AZ: Arizona
AR: Arkansas
CA: California
CO: Colorado
CT: Connecticut
DE: Delaware
DC: District of Columbia
FL: Florida
GA: Georgia
HI: Hawaii
ID: Idaho
IL: Illinois
IN: Indiana
IA: Iowa
KS: Kansas
KY: Kentucky
LA: Louisiana
ME: Maine
MD: Maryland
MA: Massachusetts
MI: Michigan
MN: Minnesota
MS: Mississippi
MO: Missouri
MT: Montana
NE: Nebraska
NV: Nevada
NH: New Hampshire
NJ: New Jersey
NM: New Mexico
NY: New York
NC: North Carolina
ND: North Dakota
OH: Ohio
OK: Oklahoma
OR: Oregon
PA: Pennsylvania
RI: Rhode Island
SC: South Carolina
SD: South Dakota
TN: Tennessee
TX: Texas
UT: Utah
VT: Vermont
VA: Virginia
WA: Washington
WV: West Virginia
WI: Wisconsin
WY: Wyoming

## 8. Declarations

### 8.1 Ethics approval and consent to participate

This study utilized publicly available, de-identified data from the Global Burden of Disease (GBD) study. As the data do not involve direct human participation and are anonymized, institutional ethics approval and informed consent were not required. The study was conducted in accordance with the principles of the Declaration of Helsinki.

### 8.2 Consent for publication

Not applicable.

### 8.3 Authors’ contributions

**Conceptualization:** Alireza Sadeghi; **Data curation:** Alireza Sadeghi, Niloofar Dehdari Ebrahimi, Ehsan Taherifard; **Formal analysis:** Alireza Sadeghi; **Investigation:** Alireza Sadeghi, Fatemeh Nouri, Wassim Kassouf; **Methodology:** Alireza Sadeghi; **Software:** Alireza Sadeghi; **Visualization:** Alireza Sadeghi; **Writing – review & editing:** Alireza Sadeghi, Fatemeh Nouri, Niloofar Dehdari Ebrahimi, Ehsan Taherifard, Mohammadreza Soltani, Stephen B. Williams, Wassim Kassouf; **Writing – original draft:** Alireza Sadeghi, Fatemeh Nouri; **Resources:** Ehsan Taherifard; **Supervision:** Mohammadreza Soltani, Stephen B. Williams, Wassim Kassouf; **Project administration:** Wassim Kassouf;

## 8.4 Acknowledgments

None.

## 8.5 Transparency Statement

The lead author, Alireza Sadeghi, affirms that this manuscript is an honest, accurate, and transparent account of the study being reported; that no important aspects of the study have been omitted; and that any discrepancies from the study as planned (and, if relevant, registered) have been explained.

